# Potential environmental transmission routes of SARS-CoV-2 inside a large meat processing plant experiencing COVID-19 clusters

**DOI:** 10.1101/2021.06.20.21259212

**Authors:** Myrna M.T. de Rooij, Reina S. Sikkema, Martijn Bouwknegt, Yvette de Geus, Kamelia R. Stanoeva, Sigrid Nieuwenweg, A.S.G. (Sandra) van Dam, Ceder Raben, Wietske Dohmen, Dick Heederik, Chantal Reusken, Adam Meijer, Marion P.G. Koopmans, Eelco Franz, Lidwien A.M. Smit

**Author notes:** corresponding author, (Institute for Risk Assessment Sciences, Utrecht University, Yalelaan 2, 3584CM Utrecht, the Netherlands;). **Supplemental Information available online** Supplemental Methods.

## Abstract

Worldwide exceptionally many COVID-19 clusters were observed in meat processing plants. Many contributing factors, promoting transmission, were suggested, including climate conditions in cooled production rooms favorable for environmental transmission but actual sampling studies are lacking. We aimed to assess SARS-CoV-2 contamination of air and surfaces to gain insight in potential environmental transmission in a large Dutch meat processing plant experiencing COVID-19 clusters.

We performed SARS-CoV-2 screening of workers operating in cooled production rooms and intensive environmental sampling during a two-week study period in June 2020. Sampling of air (both stationary and personal), settling dust, ventilation systems, and sewage was performed. Swabs were collected from high-touch surfaces and workers’ hands. Screening of workers was done using oronasopharyngeal swabs. Samples were tested for presence of SARS-CoV-2 RNA by RT-qPCR.

Of the 76 (predominantly asymptomatic) workers tested, 27 (35.5%) were SARS-CoV-2 RNA positive with modest to low viral loads (Ct≥29.7). In total, 6 out of 203 surface swabs were positive (Ct ≥38), being swabs taken from communal touchscreens/handles. One of the 12 personal air samples and one of the 4 sewage samples were positive, RNA levels were low (Ct≥38). All other environmental samples tested negative.

Although one-third of workers tested SARS-CoV-2 RT-PCR positive, environmental contamination was limited. Hence widespread transmission of SARS-CoV-2 via air and surfaces was considered unlikely within this plant at the time of investigation in the context of strict COVID-19 control measures in place.

## Introduction

Clusters of human SARS-CoV-2 infections (COVID-19) have been observed worldwide in a variety of private, public and occupational settings. Not only workers in healthcare but also workers in other essential services/industries like the food producing industry face an increased risk^1^. Exceptionally many SARS-CoV-2 outbreaks were reported in meat processing plants across Europe, Australia and the Americas^1–5^. In some cases, facilities were closed due to the high number of infected workers which was regarded as a last resort by local authorities given the necessity of food production.

A combination of several factors may explain why meat processing plants were found to be SARS-CoV-2 infection hotspots, including operational practices (e.g. high density of workers, enhanced breathing and yelling due to the physically intense work and noisy environment), societal and/or economic factors (e.g. migrant workers sharing housing and transportation), and the climate conditions inside the production rooms^1–3,6^. The low temperature, which is in place to ensure food safety, combined with presence of air recirculation systems to reduce energy use, are suggested to be advantageous for persistence and circulation of SARS-CoV-2 in air. The probable relevance of climate conditions was highlighted in experimental studies on factors affecting viability of aerosolized SARS-CoV-2^7–12^ and was suggested to play a role in an outbreak at a meat processing plant in Germany^13^ where no environmental sampling was conducted. Besides low temperatures being potentially advantageous for airborne transmission, it might also facilitate fomite transmission (touching a contaminated surface and then transferring virus to facial mucosa) as experiments showed prolonged viability of SARS-CoV-2 on surfaces with cooler temperatures^14^. However, studies including environmental sampling in meat processing plants to assess potential transmission via air and surfaces have thus far not been performed.

In the Netherlands, an increased incidence of SARS-CoV-2 infections was notified amongst workers in cooled production rooms of a high-throughput pig meat processing plant by the end of May 2020. Immediately, the COVID-19 policy of the slaughterhouse already in place was sharpened with stricter measures and supervision on compliance was intensified. In June 2020, we conducted a study to assess the role of environmental transmission of SARS-CoV-2 in this plant. The objectives were to assess potential transmission via air and surfaces. Therefore, extensive environmental sampling was performed simultaneously with voluntary screening for SARS-CoV-2 RNA in oro-nasopharyngeal swabs collected from employees.

## Methods

Details and pictures of study setting, sampling methods, and laboratory procedures are provided in the Supplemental Material.

### Investigated slaughterhouse

Investigations were performed at a high-throughput pig slaughterhouse in the Netherlands. The production process can be divided into two parts: i) process from live animals until halved carcasses, and ii) process where carcasses are further sectioned, processed and packed. The latter is performed in two large cooled production rooms (temperature: 5-9°C): a cutting room of 9,000 m^3^ and deboning room with a packaging area of 10,800 m^3^. The number of persons working in the abattoir during each shift is around 850, of whom 600 are working in cooled production rooms (215 in cutting room, 385 in deboning room/packaging area). Cooled production rooms are ventilated by a system comprising of two-stage filtering and air is largely recirculated. Each day after production, a rigorous multi-stage cleaning procedure is followed involving wetting from bottom-to-top with a mix of cleaning/disinfecting agents including chlorine-based agents.

Screening for SARS-CoV-2 RT-PCR status amongst a random selection of voluntarily participating abattoir workers on May 29^th^, showed a prevalence that was especially high among workers operating in cooled production rooms: 41% in the cutting room (9/22), 32% in the deboning room (6/19) and 16% in the packaging area (3/19) versus 0% (0/45) in other sections. From March 2020, initial COVID-19 measures were implemented involving prevention of close contact between workers (separation of work shifts and breaks in time, work place modifications) and increased focus on hand hygiene at entry of the premises and in non-production locations. From the start of June, additional measures were implemented involving intensified cleaning and disinfection procedures (incl. air treatment by fogging every Sunday with hydrogen peroxide and lactic acids), a triage based on symptoms (questionnaire and interview) of all individuals entering and contact reductions while commuting.

### Sampling strategy

Environmental sampling was performed at three time-points in June 2020 (T1: June 8, T2: June 15, T3: June 19). SARS-CoV-2 RT-PCR screening of a random selection of workers by oro-nasopharyngeal sampling was performed at T2, and screening based on sewage sampling at T1 and T2. To assess potential transmission via air, we performed sampling of air, settling dust and filters of the ventilation system. To assess potential transmission via surfaces, swabs were collected from surfaces that were expected to be touched frequently as well as the hands/gloves of workers. At T1 the purpose of environmental sampling was to gain broad insight into potential environmental SARS-CoV-2 RNA presence in the various areas either in air or on surfaces. Stationary air sampling was performed at potential hotspots based on workers’ density and ventilation characteristics in both production rooms. Environmental swabs were used to sample a selection of various high-touch surfaces present throughout the facility. At T2, focus was on personal air sampling during the shift of workers participating in SARS-CoV-2 oro-nasopharyngeal screening combined with swabbing of their hands/gloves enabling. Environmental swabs were taken from high-touch surfaces not yet sampled. At T3, environmental swabs were collected from same and similar high-touch surfaces identified to be relevant at T2. Throughout the study, strict safety and hygienic procedures were followed to prevent infection and contamination. Field blanks of all sample types were collected as a control.

### Screening and scoring

Sewage samples (2 tubes of 50ml 24-hour flow dependent composite sample) were collected as described previously^15^ at both T1 and T2 in collaboration with the external water treatment plant located at the facility. At T2, in collaboration with the municipal health services (GGD), oro-nasopharyngeal swabs were collected from persons working at the cooled production rooms before and after the shift (minimum working time: 6.5 hours). Questionnaires were collected including items on health status, contacts and working and living conditions. Workers participated on a voluntary basis, informed consents were obtained. Each worker received 40 euros for participation.

Workers were scored on SARS-CoV-2 transmission relevant behaviour and personal protective measures (PPM) by means of scoring cards by fieldworkers. To gain an overall impression of wearing surgical masks (categorized: covering nose and mouth, covering mouth, or not-wearing), a minimum of 45 persons in both production rooms were scored. In addition, 5-minute observations of workers performing their job-tasks were performed to note wearing of PPM and physical distancing (both for longer durations, e.g. conversations, and solely passing).

### Sampling air and surfaces

Air sampling methodology was similar as described previously by De Rooij et al^16^. In short, a filter-based technique was used to sample inhalable dust—airborne particles small enough to enter the respiratory tract. For stationary air sampling, sampling heads were attached onto a pole at 1.50m height (average breathing height of humans). Personal air sampling was performed by attaching the sampling head within the breathing zone of the worker. Stationary 6-hour sampling was performed in both production rooms. At T1, sampling was performed at 5 sites per room. At T2, stationary sampling was performed at 2 sites per room; the remainder of sampling equipment was used for personal sampling. Of the workers participating in oro-nasopharyngeal screening, 12 workers (6 per room) were selected to participate in personal air sampling. Personal air sampling was performed from the beginning until the end of the worker’s shift, resulting in 6 to 8 hour measurements.

Sampling of settling dust in production rooms and the canteen was performed by using Electrostatic Dust fall Collectors (EDCs), which contain electrostatic cloths placed in a disposable holder, as described previously^17^.

Sampling of the ventilation system was performed at T2 for both production rooms. Per room, one filter of each type (Coarse 50% and ePM10 80%/ePM2.5 70%) was collected from their respective grid. These filters had been placed in August 2019.

Swabs of high-touch surfaces were collected in the production rooms and in all other areas workers have access to (e.g. canteen area, locker room, toilets). Per time-point, at least 60 surface swabs were taken throughout these areas. Swabs of hands, or gloves if worn, of the 12 workers participating in the personal air sampling were collected during their mid-shift break.

### Sample processing and laboratory procedures

Samples were stored after collection at 4°C until further processing within 24-hours at BSL-2 conditions. Total nucleic acid was extracted from oro-nasopharyngeal samples using a MagNA Pure 96 with total nucleic acid small volume kit (Roche). Thereafter, samples were tested for the presence of SARS-CoV-2 RNA using RT-qPCR, targeting the E-gene and the RdRP-gene with detection limits at 3.2 and 3.7 RNA copies/reaction respectively^17^. A worker was defined positive if at least one of the two genome targets tested positive in one or both swabs.

On the other samples, RNA extraction was performed using an in-house method using Ampure beads^18^. These samples were tested for the presence of SARS-CoV-2 RNA using RT-qPCR, targeting the E-gene (detection limit 3.3 RNA copies/reaction)^19,20^.

## Results

### Screening

Of the 81 workers invited, 76 (94%) participated in the oro-nasopharyngeal SARS-CoV-2 screening performed at T2. One worker solely participated in the pre-shift sampling round (sample tested negative). In total, 27 workers (35.5%) tested positive for SARS-CoV-2 RNA (Table 1). Of the cutting room workers, 21% tested positive versus 50% of the deboning area workers. Most workers were Polish or Romanian, in both groups 40% tested positive. For 6 persons (22% of the test-positive cases) SARS-CoV-2 RNA was detected in both pre-and post-shift swabs. Seventeen workers tested positive pre-shift and negative post-shift, while only 4 workers tested negative pre-shift and positive post-shift. Ct-values ranged between 29.7 and 38.3 for E-gene and between 31.2 and 39.6 for RdRp-gene (Figure 1), corresponding to modest to low viral loads. Of the 76 workers, 74 (97%) filled in the questionnaire. The two workers that did not return the questionnaire tested SARS-CoV-2 negative. None of the surveyed employees classified themselves as symptomatic at entrance triage. However, three testing-negative and two testing-positive workers did report mild symptoms in our questionnaire (Table 1). At T2, one sewage sample tested positive (Ct-value 39 corresponding to approx. 5.5 copies/ml sewage).

**Table 1.** Characteristics of 76 meat processing workers participating in naso-oropharyngeal SARS-CoV-2 RNA screening performed on June 15^th^ 2020

|  | N | SARS-CoV-2<br>negative<br>N=49 (64.5%) | SARS-CoV-2<br>positive<br>N=27 (35.5%) |
| --- | --- | --- | --- |
| **Cooled production room** |  |  |  |
| Cutting | 38 | 30 (79%) | 8 (21%) |
| Deboning | 38 | 19 (50%) | 19 (50%) |
| **Nationality** |  |  |  |
| Hungarian | 5 | 5 (100%) | 0 (0%) |
| Lithuanian | 1 | 1 (100%) | 0 (0%) |
| Polish | 15 | 9 (60%) | 6 (40%) |
| Portuguese | 1 | 1 (100%) | 0 (0%) |
| Romanian | 53 | 32 (60%) | 21 (40%) |
| Slovak | 1 | 1 (100%) | 0 (0%) |
| **Current residential situation** |  |  |  |
| Alone | 13 | 8 (62%) | 5 (38%) |
| Shared with up to 4 housemates | 41 | 26 (63%) | 15 (37%) |
| Shared with 5 or more housemates | 19 | 13 (68%) | 6 (32%) |
| **Mode of transportation to work** |  |  |  |
| Alone (car/bike) | 29 | 20 (69%) | 9 (31%) |
| By public transport | 3 | 2 (67%) | 1 (33%) |
| By car/mini-van with other people | 42 | 25 (60%) | 17 (40%) |
| **Current province of residence** |  |  |  |
| Noord-Brabant (NL) | 68 | 44 (65%) | 24 (35%) |
| Nordrhein-Westfalen (DE) | 8 | 5 (63%) | 3 (37%) |
| **COVID-19 related symptoms<sup>a</sup> (n=74)** |  |  |  |
| Without symptoms (self-reported) | 69 | 44 (64%) | 25 (36%) |
| With symptoms (self-reported) | 5 | 3 (60%) | 2 (40%) |
| **Chronic disease status<sup>b</sup> (n=74)** |  |  |  |
| Without chronic condition (self-reported) | 71 | 45 (63%) | 26 (37%) |
| With chronic condition (self-reported) | 3 | 2 (66%) | 1 (33%) |

Note. Per characteristic, the number of participants is noted for which this data was available.
<sup>a</sup> Self-reported potential COVID-19 related symptoms included runny nose and loss of smell and/or taste (1 worker test-positive and 1 worker test-negative), fever or feeling warm and loss of smell and/or taste (1 worker test-negative), headache (1 worker test-negative), having a cough maybe/don't know (1 worker test-positive) .
<sup>b</sup> Chronic disease status defined as positive answer to question 'Do you have a chronic disease'. The person with chronic disease testing SARS-CoV-2 RNA positive reported hypertension controlled by beta blockers.

**Figure 1.**
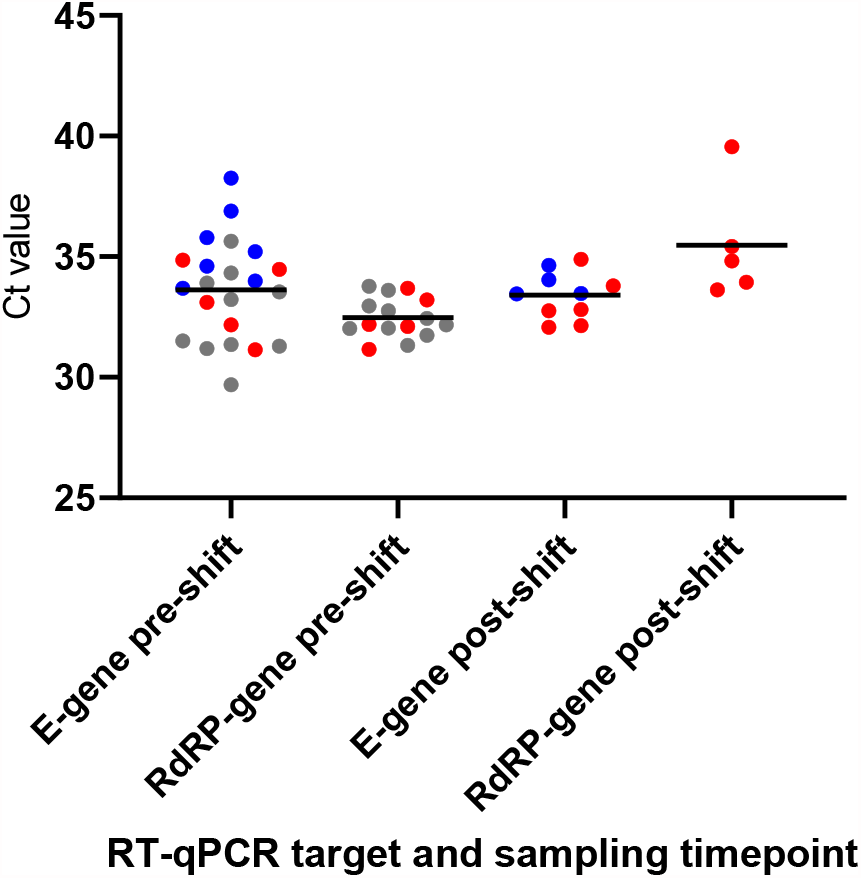
Distribution of Ct-values by gene target and moment of sampling (pre-shift, post-shift) detected in oro-nasopharyngeal swabs from 27 meat processing workers tested SARS-CoV-2 RNA positive. Note. Red dots indicate six employees that were positive at both sampling moments (pre-shift and post-shift) for one or two target genes; blue dots indicate eleven employees who were positive for one target gene and one sampling moment; grey dots indicate ten employees who were positive for both target genes pre-shift only. The horizontal bar indicates the mean Ct-value.

### Air and surfaces

In total 271 samples were collected (Table 2). At T2, SARS-CoV-2 RNA was detected in 9.8% of the surface swabs (6/61, Ct-values 38 to 39 corresponding to approx. 8×10^1^ to 1.6×10^2^ copies per swabbed surface). Of the 22 surface swabs collected at the cutting room at T2, three (14%) swabs tested positive, taken from a machine handle (with ridges), grip side of a stepladder, and the handle of a pressure pump used for disinfection. Of the 18 surface swabs collected at non-production areas at T2, three (17%) tested positive: swabs taken from a touch screen on the coffee machine, main touch screen for lockers in a changing room, and handle of a dispenser used for hand disinfecting. All 6 positive surfaces can be classified as high-touch. All 21 surface swabs collected in the deboning room at T2 were negative. All 142 surface swabs collected at T1 and T3 in production rooms as well as non-production areas were negative.

**Table 2.** SARS-CoV-2 PCR test results of in total 275 samples taken of air, surfaces, workers’ hands and sewage in a meat processing plant

| Sampling time-point | Sample type | % positive samples (n positives/N) |
| --- | --- | --- |
| T1 | Inhalable dust - stationary | 0% (0/10) |
| T1 | EDC | 0% (0/16) |
| T1 | Surface swab | 0% (0/68) |
| T1 | Sewage water | 0% (0/2) |
| T2 | Inhalable dust - stationary | 0% (0/4) |
| T2 | Inhalable dust - personal | 8.3% (1/12) |
| T2 | EDC | 0% (0/6) |
| T2 | Surface swab | 9.8% (6/61) |
| T2 | Sewage water | 50% (1/2) |
| T2 | Ventilation system filter | 0% (0/8) |
| T2 | Swab of hand worker | 0% (0/12) |
| T3 | Surface swab | 0% (0/74) |

SARS-CoV-2 RNA was detected in one of the 12 personal air samples (Ct-value 38 corresponding to approx. 5×10^2^ copies/m^3^). The worker with the SARS-CoV-2 positive air sample, tested oro-nasopharyngeal positive at the start of the shift (Ct-value 33.2 E-gene, 33.8 RdRp-gene), but tested negative post-shift. Of the other 11 workers participating in the personal air sampling, one worker had a positive pre-shift and post-shift test (Ct-value E-gene 34.9, 32.8, respectively; RdRp-gene 33.7, 33.6); five workers only had a positive pre-shift swab (range in Ct-values E-gene 33.5-35.6; RdRp-gene 31.7-33.6 and two >40). SARS-CoV-2 RNA was not detected in any of the stationary inhalable dust samples (T1, n=10; T2, n=4). All other sample types (settling dust, filters ventilation system, swabs of workers’ hands) also tested negative.

### Observations

The majority of the 100 scored workers wore a surgical mask covering solely the mouth (66%, 29/40 cutting workers; 75%, 30/40 deboning workers; 55%, 11/20 packaging workers), others wore the mask covering mouth and nose. One person (deboning area) did not wear a mask. All of the 12 personal air sampling participants wore a mask, 11 (92%) wore the mask covering solely the mouth. Of the 11 personal air sampling participants with a negative air sample, 9 had a stationary job task and few persons passed by their fixed positions along the line (most kept 1.5m distance). Seven of them worked at a position with 8 or more persons working in 10m vicinity, the other two workers were surrounded by respectively 2 and 4 persons. The 2 workers with non-stationary tasks, showed frequent passing-by or being passed-by within 1.5m distance (several times per minute). The only worker with a positive personal air sample had a stationary job task in the deboning room and was surrounded by 10 persons in 10m vicinity with a distance of >1.5m from the nearest worker. Observations of personal air sampling participants were similar to 10 randomly selected workers per production room with respect to surrounding workers and 1.5m distancing.

## Discussion

Our findings provide compelling leads to the relevance of environmental transmission of SARS-CoV-2 in a large meat processing plant. SARS-CoV-2 RNA was detected in one personal air sample, and on six frequently touched surfaces. Screening of workers’ SARS-CoV-2 status by oro-nasopharyngeal swabbing showed a considerable percentage of workers to be SARS-CoV-2 RNA positive, with a relatively low viral load and generally without symptoms. Results of environmental sampling showed a low number of SARS-CoV-2 RNA positive samples overall which suggests a limited role of transmission via air and surfaces inside the cooled production rooms during the two-week study period. Results should be interpreted in the context of strict prevention and mitigation measures in place at the time the study was performed.

### SARS-CoV-2 status of workers

Our investigation showed that one third of the tested workers were positive for SARS-CoV-2 RNA in at least one of the two oro-nasopharyngeal swabs collected pre-and post-shift. Viral loads detected in the swabs were low and workers were predominantly asymptomatic. There are several hypotheses to explain these findings: i) worker(s) may have experienced a (mild) infection in the past without noticing/recalling symptoms (post-infection scenario), ii) worker(s) could be in pre-symptomatic state at the time of sampling (pre-symptomatic scenario), iii) worker(s) could experience an asymptomatic infection (asymptomatic scenario). Published meta-analyses reported percentages of SARS-CoV-2 infected persons remaining asymptomatic throughout infection of around 15-20%^21–23^, although percentages can be higher in single-family clusters (95% CI: 26%– 44%)^22^. SARS-CoV-2 RNA can remain detectable in swabs from the upper respiratory tract several weeks to months after onset of infection^24,25^. As workers that tested positive were followed-up and no clear symptoms suggestive of COVID-19 had developed, the pre-symptomatic scenario seems unlikely leaving both scenarios of post-infection and asymptomatic as realistic. If we consider low RNA loads in participating workers a proxy of viral excretion^25–27^, high shedding rates of infectious SARS-CoV-2 are not to be expected. The majority of workers tested positive only pre-shift, which may be explained by physiological accumulation of respiratory tract secretions at the start of the day^28^, swabbing differences between testers^29^, and/or influence of stochastic processes especially at low viral loads (higher chance of false-negatives). SARS-CoV-2 RNA level in the positive sewage sample was comparable to levels detected at urban sewage sites in the Netherlands in the early stage of the epidemic (March 2020)^30^. Because of site-to-site dissimilarities and methodological differences^30,31^, exact prevalence cannot be estimated but points to a limited number of acute infections.

### Exposure assessment and risk estimation

Findings indicated absence of considerable SARS-CoV-2 levels in air throughout the cooled production areas. None of the stationary air samples were positive, despite the selection of likely hotspots. Central ventilation system filters were also all negative while it has been suggested that SARS-CoV-2 RNA may accumulate in filters^32^. One of 12 personal air samples was positive, with a 100-fold lower level than personal exposure levels measured in SARS-CoV-2 infected mink farms^16^. As the Ct-value of this air sample was too high for whole genome sequencing, and this worker’s oro-nasopharyngeal swab tested positive, it could not be determined whether SARS-CoV-2 RNA detected in this personal air sample originated from this individual, and/or from other workers. Low or non-detectable exposure as found in personal air samples can be explained by COVID-19 measures in place^33^ (e.g. physical distancing, masks) and limited viral shedding by workers in line with low viral loads in oro-nasopharyngeal screening and negative personal air samples for 6 positive-tested workers. Inhalation exposure during a workday to such low/non detectable levels of SARS-CoV-2 RNA (and even lower levels of viable virus), is not expected to pose a high risk of infection^34^. Deposition of inhaled SARS-CoV-2 contaminated particles anywhere along the respiratory tract, from nasal epithelial cells to deep in the airways, has the potential to initiate infection^35^ so air sampling covered the relevant particle size fraction. As no viability testing was performed, no inferences on potential levels of viable virus could be made.

The many surfaces sampled showed limited SARS-CoV-2 surface contamination, with low viral RNA loads in a few positive samples. As the hygiene standards in the food processing industry are high^36,37^, regulations are already in place to ensure frequent and proper hand washing and disinfecting. This was substantiated by swabs from workers’ hands/gloves being all negative for SARS-CoV-2 RNA. Considering limited SARS-CoV-2 RNA surface contamination observed (thus even lower considering viable virus), and focus on hand hygiene is in place, we consider this not a main route of transmission in this meat processing plant during the study period. Given the sampling design (focus on major high-touch surfaces, sampling later during the day so both shifts have passed), it is unlikely that the level of surface contamination at the time of investigation was underestimated. Pork carcasses or meat products as a possible source can be excluded, as animal studies showed that pigs are unlikely to get infected with SARS-CoV-2^38,39^.

### Comparisons to other research in meat processing plants

The importance of the airborne route has been suggested in particular by an outbreak investigation performed in a large meat processing plant in Germany, which did not look at the potential role of surface contamination^13^. Based on spatio-temporal aspects of the outbreak, it was suggested that SARS-CoV-2 had efficiently spread inside the production room via distances of more than 8 meters but no environmental sampling was performed. Airborne transmission was not supported by our air sampling, but differences between facilities (e.g. lay-out, ventilation system and air flow), COVID-19 incidence, and control measures in place at the time of investigation preclude firm conclusions on the airborne route. Research performed in the United States on evaluation of effectiveness of COVID-19 measures in meat processing plants suggested mitigation of transmission after initiating universal mask policy and installing physical barriers but the exact effect could not be determined as not all, potentially confounding, factors (e.g. other measures implemented, transmission beyond the workplace) could be assessed^40^.

### Considerations on COVID-19 policy

At the time the study was performed, strict preventive and mitigation measures were already in place, so effectiveness of individual interventions can only be speculated upon. Even more intense cleaning could be recommended for exceptionally high-touch surfaces in the non-production rooms (touchscreens and handle) and non-smooth surfaces in the production rooms (handles/grip side). Based on our findings, the meat processing plant decided to further intensify cleaning and disinfection of these surfaces during the day. Entrance triage appeared not completely effective in preventing persons with potential COVID-19 related symptoms going to work emphasizing, on top of the risk of asymptomatic infections, the importance of mitigation measures. Physical distancing measures, which were mostly adhered to according to observations of workers’ behaviour, may have limited inhalation exposure. The effectiveness of surgical masks to prevent shedding by positive persons and protect susceptible persons cannot be properly estimated as many influential factors are suggested including mask-related (e.g. type and fit) and human-related factors (e.g. way of wearing, use in general including donning, doffing, renewal)^33,41,42^. In cooled production rooms, standard surgical masks can cause discomfort/annoyance as glasses fog easily and masks typically become moist quickly, which also may deteriorate effectiveness^43^. Studies on PPE including mask wearing are highly warranted in occupational settings like these, to evaluate effectiveness and user-friendliness in practice and to provide concrete evidence-based advise on policy. Proper ventilation and cleaning of the ventilation system could have contributed to low virus levels in the air^44^, but effectiveness of ventilation in reducing SARS-CoV-2 transmission remains to be quantified.

To conclude, given the overall low number of environmental samples positive for SARS-CoV-2 RNA, widespread transmission of SARS-CoV-2 via air and surfaces within this meat processing plant was not considered likely at the time of investigation, and could be a consequence of the many COVID-19 control measures in place. Studies evaluating interventions in real-life settings are highly warranted to better understand the role of environmental transmission of SARS-CoV-2, and to guide proper control measures to take in occupational settings.

## Data Availability

The datasets generated during and/or analysed during the current study are available from the corresponding author on reasonable request.

## Acknowledgements

We gratefully acknowledge all persons contributing to sample collection and/or in the lab: Thijs Groeneveld, Isabella van Schothorst, Jack Spithoven, Aniek Lotterman, Inge Wouters, Pascalle Roulaux, Calvin Ge, Olivier Horstink, Derk Oorburg, Bart de Ruiter, Irina Chestakova, Anne van der Linden, Gabriel Goderski, Lisa Wijsman, Sharon van den Brink and Harry Vennema. We gratefully thank the participating slaughterhouse employees for their cooperation.

## Ethics Approval statement

The Medical Research Ethics Committee (MREC) Utrecht confirmed that the Medical Research Involving Human Subjects Act (WMO) did not apply to this study, and that therefore an official approval of this study by the MREC Utrecht was not required under the WMO (Protocol 20-385/C, reference number WAG/mb/20/021975).

## Supplemental Methods

### Additional information on the investigated meat processing plant

The abattoir is in production six days a week (Monday-Saturday) and per day two consecutive shifts are scheduled (morning shift and afternoon/evening shift, with the exception of Saturday with solely a morning shift). In general, workers are scheduled to work one week in the morning shift and the next week in the afternoon shift in pools with stable composition. Workers typically have a fixed job task and operate at the same position along the processing line. There is a strict separation between the first (non-cooled) and second part (cooled) of the production process regarding personnel, areas accessible to personnel, materials and clothing.

Cooled production-associated areas are solely accessible for workers operating in the cooled production rooms. These include a canteen area with restaurant, various changing rooms with lockers and toilet facilities, passageways with staircases and one large hygiene lock. Areas are cleaned daily and toilet facilities cleaned twice a day with designated cleaning/disinfecting agents including chlorine-based agents.

Rooms are ventilated by a system comprising of two-stage filtering. The first stage includes a filter for larger particles (ISO 16890 Coarse 50%), the second stage includes a filter for smaller particles (ISO 16890 ePM_10_ 80% and ISO 16890 ePM_2.5_ 70%). Air is largely being recirculated, with minimally passive air refreshment through *e*.*g*. open inner doorways and corridors. Rooms are being thoroughly cleaned each day after production. A rigorous multi-stage procedure is followed involving wetting from bottom until top with a mix of cleaning/disinfecting agents including chlorine-based agents. Since June 2020, fogging was also performed each Sunday with hydrogen peroxide and lactic acid as active substances.

Face shields were solely worn by workers with communication duties (e.g. foremen/intendents) in line with the slaughterhouse’s policy.

### Additional information on sampling

#### Oro-nasopharyngeal sampling

Oro-nasopharyngeal sampling was performed according to the June 2020 prevailing national monitoring protocol (https://lci.rivm.nl/richtlijnen/covid-19); the same swab was used to first swab the oropharynx followed by the nasopharynx. Thereafter, the swab was directly placed in 3 ml GLY virus transport medium.

#### Questionnaires

Self-reported data obtained from employees was collected using questionnaires focusing on the following topics: current and prior symptoms, overall health and chronic conditions, living situation, contact with workers in meat-processing facilities, contact with COVID-19 cases, recent travel history, work situation and workplace, contact with co-workers, and commuting. The questionnaires were available in five languages (Dutch, English, Polish, Hungarian and Romanian). Questionnaires were distributed upon the first sampling and collected upon the second sampling.

#### Personal and stationary air sampling

Teflon filters (Pall Corporation, Ann Arbor, USA) were used in GSP (Gesamtstaubprobenahme, total dust sampling; JS Holdings, Stevenage, UK) sampling heads connected to a Gilian GilAir 5 pump (Sensidyne, St. Petersburg, USA) calibrated at a flow of 3.5 l/min.

Picture showing stationary air sampling (orange circle around sampling head):

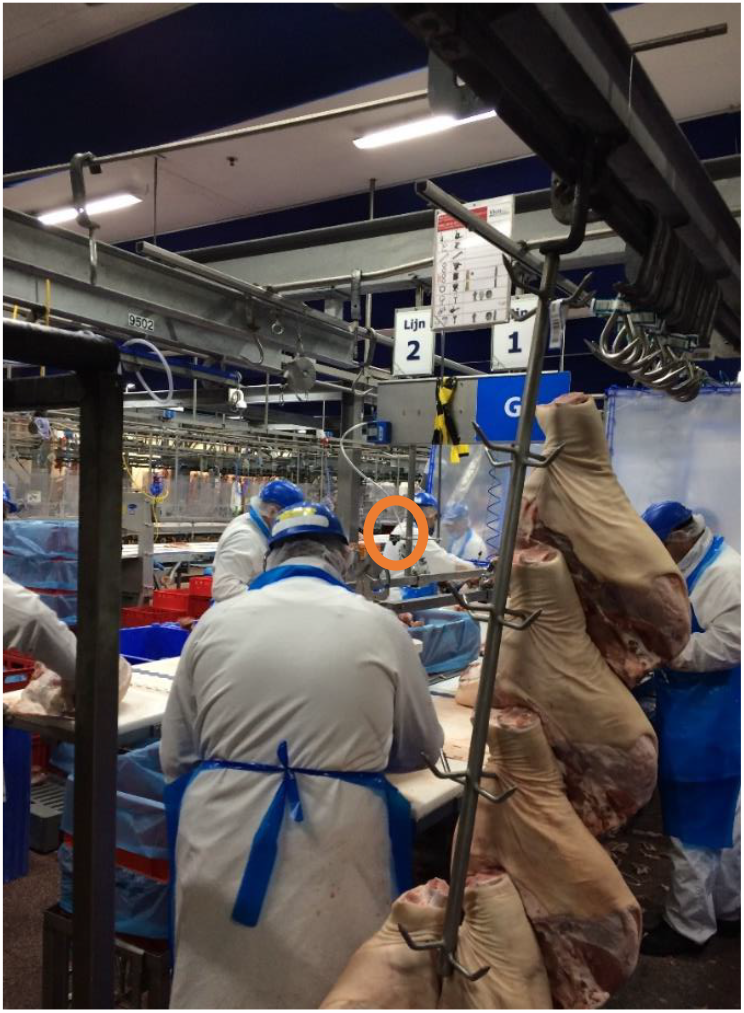

Pictures showing personal air sampling:

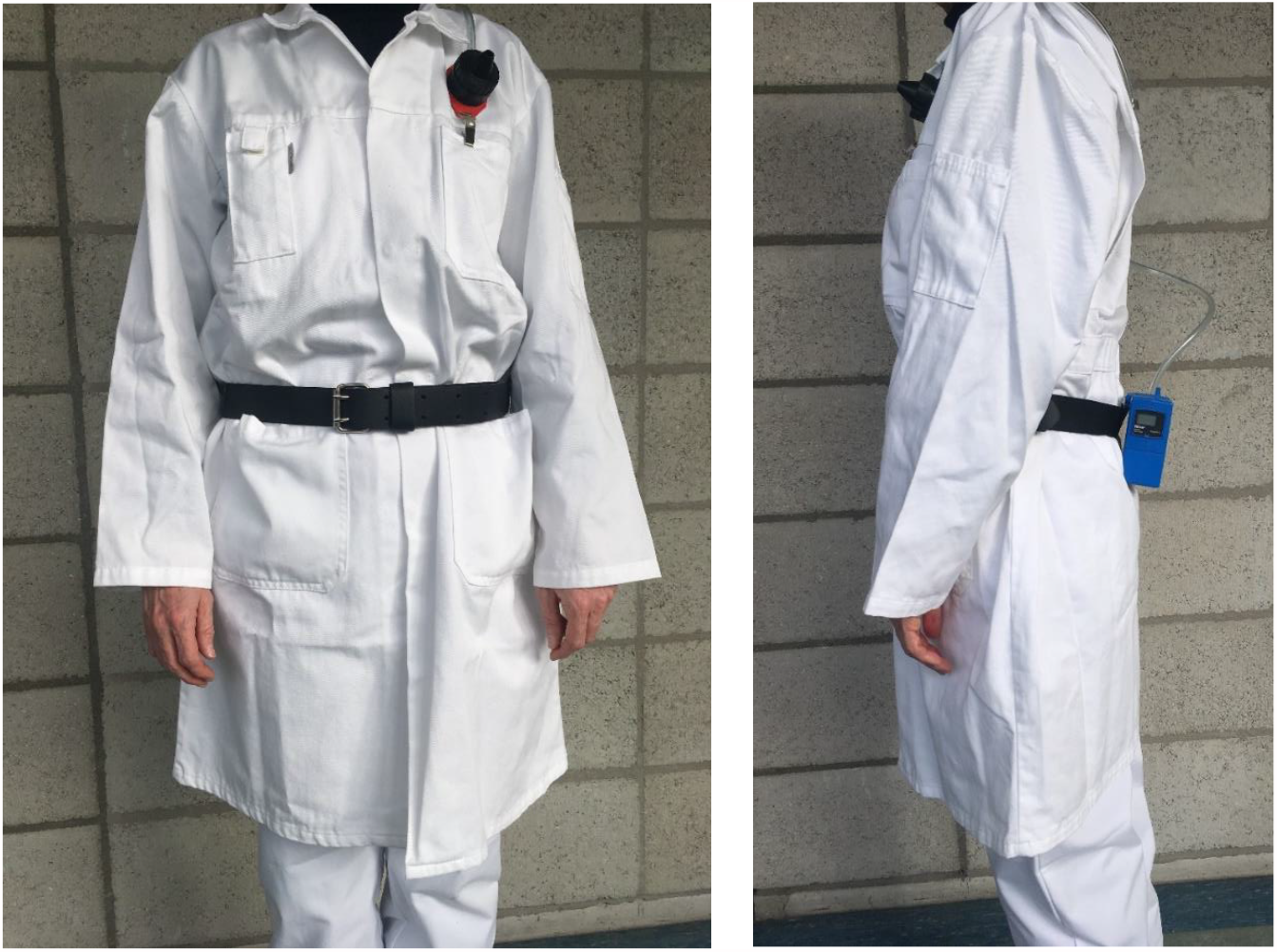

Note. Due to privacy reasons, a picture of a researcher was taken and not of a slaughterhouse worker

#### Electrostatic Dust fall Collectors (EDCs)

EDCs are sterilized electrostatic cloths (polyester electrostatic cloth; Albert Heijn, Zaandam, the Netherlands) placed in a disposable holder. At T1, 5 EDCs were placed per production room; these could only be exposed during one day due to the cleaning regime involving bottom-to-top wetting. EDCs placed in the canteen area were exposed during 7 days. At T1, 6 EDCs were placed in the canteen area, which were collected at T2 and replaced with new EDCs which were collected at T3.

Picture showing EDC positioned on top of machine in cutting room:

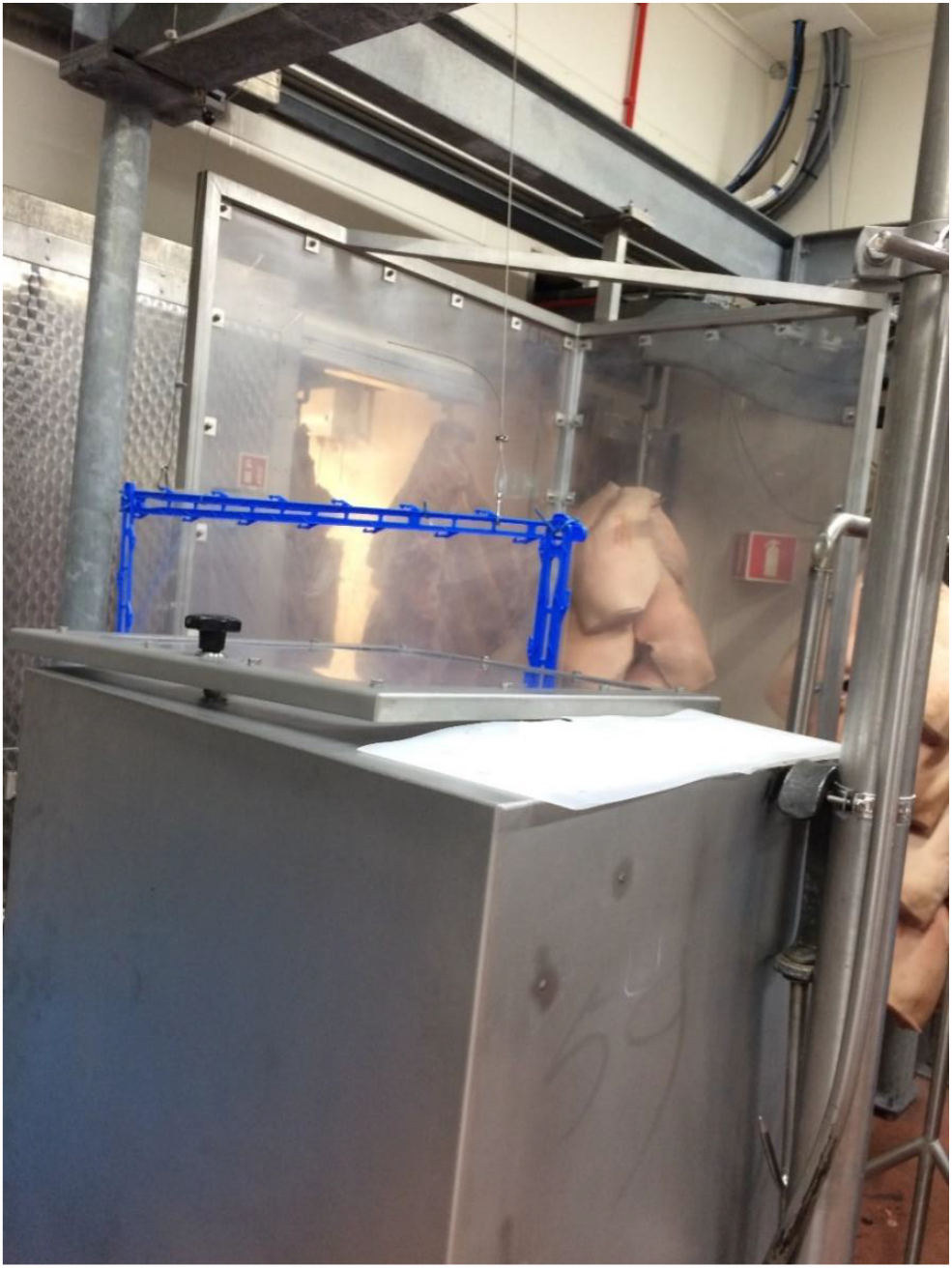

#### Surface swabs

Swabbed items included knobs, grips, push buttons, touchscreens and all sorts of handles (e.g. machinery in production rooms, dispensers in toilets); but also table tops, chairs, stair railings and other surfaces frequently touched. Disposable plastic grids of 10 cm^2^ were used for standardization of the sampled surface. If a surface was smaller than 10 cm^2^ this was noted. Dry swabs with a rayon tip and plastic shaft (CLASSIQSwabs 167KS01; COPAN, Brescia, Italy) were used, which were placed in 2ml virus transport medium (VTM) directly after swabbing.

#### Swabs of workers’ hands/gloves

Dry swabs with a rayon tip and plastic shaft (CLASSIQSwabs 167KS01; COPAN, Brescia, Italy) of hands, or gloves if worn, of the 12 workers participating in the personal air sampling were collected during their mid-shift break. The inner part of their index finger, middle finger and ring finger was swabbed until the half of their palm.

#### Specifications of fieldworkers and field blanks

Fieldworkers that visited the meat processing plant for sampling were routinely monitored for SARS-CoV-2 infection (remained negative throughout the study). Field blanks of all sample types were collected as a control, these blanks underwent all procedures (e.g. preparations, transportation, processing) as the actual samples except for sampling. All field blanks tested negative.

### Details on laboratory analyses

#### Electrostatic Dust fall Collectors (EDCs)

One tablet of protease inhibitor was dissolved in 20mL D-PBS, without Calcium and Magnesium. Half of the EDC/mouth mask was put into a 50mL tube, containing 10 mL D-PBS+protease inhibitor and incubated for 1 hour at room temperature on a tube roller. 60uL sample was removed, and 90 ul MagNA Pure 96 External Lysis Buffer (Roche) was added. PDV (10 uL) was used as internal control, as described previously^40^. RNA was eluted in 30 uL distilled water, 8 uL was used for the SARS-CoV-2 PCR, as described previously^17^.

#### Surface swabs and swabs of workers’ hand/gloves

Swabs in 1 ml VTM were vortexed, and 60 uL VT was further processed as described above.

#### Air filters (Teflon and ventilation system)

Teflon filters were collected from the GSP sampling heads and transferred to a 15ml tube. Per ventilation system filter, 2 punches were taken (25mm diameter, approximately 6mm thick). Each punch was transferred to a 15ml tube. To each tube 1ml VTM and 1 ml lysisbuffer (MagNA Pure 96 External, Roche) was added and subsequently vortexed for 5 minutes. PDV (10uL) was added to 150 ul of sample. RNA extraction and SARS-CoV-2 RT-PCR was performed as described above.

#### Sewage

Each tube containing 50 ml of sewage was spinned down (3000g during 15 min) and 15 ml of the supernatant was transferred to an Amicon tube. The sample was subsequently spinned down during 30 min at 4000g. The filter was rinsed with PBS and transferred into a new tube. RNA extraction and SAR-CoV-2 RT-PCR was performed as described above.

#### Schematic overview of PCR analysed proportion per sample type

| Sample type | Original sample | Volume of medium added (ml) | Volume aliquot of sample for PCR (µl) | Volume added to aliquot (µl) | Volume eluens (µl) | Volume in PCR (µl) | Proportion volume PCR/volume eluens | Proportion volume aliquot/sample volume medium | Proportion subsample of original sample |
| --- | --- | --- | --- | --- | --- | --- | --- | --- | --- |
| Teflon filter T2 (filter used in active air sampling) | 1 filter ~ cubic meter of air sampled dependent on flow rate and sampling duration | 2 ml (1 ml VTM + 1 ml lysisbuffer) | 150 → 75 µl sample | 10 µl PDV | 30 | 8 | 0.267 | 0.075 | 0.00415 |
| Swab T2 | 1 swab ~ swabbed surface of max 10 cm<sup>2</sup> | 1 ml VTM | 60 | 100 µl (90 µl lysis buffer + 10 µl PDV) | 30 | 8 | 0.267 | 0.06 | 0.01602 |
| Sewage sample T2 | 15 ml out of 50 ml tube concentrated to 200 µl ~ 24 h flow-dependent composite sample | None | 60 | 100 µl (90 µl lysis buffer + 10 µl PDV) | 30 | 8 | 0.267 | 0.3 | 0.0801 |

## References

1. Carlsten C, Gulati M, Hines S, et al. COVID-19 as an occupational disease. Am J Ind Med. 2021;(September 2020):1–11. doi:10.1002/ajim.23222

2. Middleton J, Reintjes R, Lopes H. Meat plants—a new front line in the covid-19 pandemic. BMJ. 2020;370:m2716. doi:https://doi.org/10.1136/bmj.m2716

3. Dyal JW, Grant MP, Broadwater K, et al. COVID-19 Among Workers in Meat and Poultry Processing Facilities ? 19 States, April 2020. MMWR Morb Mortal Wkly Rep. 2020;69(18):557–561. doi:10.15585/mmwr.mm6918e3

4. Steinberg J, Kennedy E, Basler C, et al. COVID-19 Outbreak Among Employees at a Meat Processing Facility - South Dakota. MMWR Morb Mortal Wkly Rep. 2020:69(31):1015–1019.

5. Waltenburg M, Victoroff T, Rose C, Butterfield M, Jervis R, Fedak M. COVID-19 Among Workers in Meat and Poultry Processing Facilities - United States. MMWR Morb Mortal Wkly Rep. 2020:69(27):887–892.

6. Taylor CA, Boulos C, Almond D. Livestock plants and COVID-19 transmission. Proc Natl Acad Sci. 2020;117(50):202010115. doi:10.1073/pnas.2010115117

7. Aboubakr HA, Sharafeldin TA, Goyal SM. Stability of SARS-CoV-2 and other coronaviruses in the environment and on common touch surfaces and the influence of climatic conditions: A review. Transbound Emerg Dis. Published online 2020. doi:10.1111/tbed.13707

8. Chin AWH, Chu JTS, Perera MRA, et al. Stability of SARS-CoV-2 in different environmental conditions. The Lancet Microbe. 2020;1(1):e10. doi:10.1016/s2666-5247(20)30003-3

9. Van Doremalen N, Bushmaker T, Morris DH, et al. Aerosol and surface stability of SARS-CoV-2 as compared with SARS-CoV-1. N Engl J Med. 2020;382(16):1564–1567. doi:10.1056/NEJMc2004973

10. Matson MJ, Yinda CK, Seifert SN, et al. Effect of environmental conditions on sars-cov-2 stability in human nasal mucus and sputum. Emerg Infect Dis. 2020;26(9):2276–2278. doi:10.3201/eid2609.202267

11. Dabisch P, Schuit M, Herzog A, et al. The influence of temperature, humidity, and simulated sunlight on the infectivity of SARS-CoV-2 in aerosols. Aerosol Sci Technol. 2021;55(2):142–153. doi:10.1080/02786826.2020.1829536

12. Morris DH, Yinda KC, Gamble A, et al. The effect of temperature and humidity on the stability of SARS-CoV-2 and other enveloped viruses. bioRxiv Prepr Serv Biol. Published online October 16, 2020. doi:10.1101/2020.10.16.341883

13. Guenther T, Czech-Sioli M, Indenbirken D, et al. Investigation of a superspreading event preceding the largest meat processing plant-related SARS-Coronavirus 2 outbreak in Germany. SSRN Electron J. Published online 2020. doi:10.2139/ssrn.3654517

14. Marquès M, Domingo JL. Contamination of inert surfaces by SARS-CoV-2: Persistence, stability and infectivity. A review. Environ Res. 2021;193(November 2020). doi:10.1016/j.envres.2020.110559

15. Izquierdo-Lara R, Elsinga G, Heijnen L, et al. Monitoring SARS-CoV-2 Circulation and Diversity through Community Wastewater Sequencing, the Netherlands and Belgium. Emerg Infect Dis. 2021;27(5):1405–1415. doi:10.3201/eid2705.204410

16. de Rooij MMT, Hakze-Van der Honing RW, Hulst MM, et al. Occupational and environmental exposure to SARS-CoV-2 in and around infected mink farms. medRxiv. Published online January 14, 2021:2021.01.06.20248760. doi:10.1101/2021.01.06.20248760

17. Noss I, Wouters IM, Visser M, et al. Evaluation of a low-cost electrostatic dust fall collector for indoor air endotoxin exposure assessment. Appl Environ Microbiol. 2008;74(18):5621–5627. doi:10.1128/AEM.00619-08

18. Richard M, Kok A, de Meulder D, et al. SARS-CoV-2 is transmitted via contact and via the air between ferrets. Nat Commun. 2020;11(1). doi:10.1038/s41467-020-17367-2

19. Iglói Z, leven M, Abdel-Karem Abou-Nouar Z, et al. Comparison of commercial realtime reverse transcription PCR assays for the detection of SARS-CoV-2. J Clin Virol. 2020;129(June):4-6. doi:10.1016/j.jcv.2020.104510

20. Corman VM, Landt O, Kaiser M, et al. Detection of 2019 novel coronavirus (2019-nCoV) by real-time RT-PCR. Eurosurveillance. 2020;25(3):2000045. doi:10.2807/1560-7917.ES.2020.25.3.2000045

21. He J, Guo Y, Mao R, Zhang J. Proportion of asymptomatic coronavirus disease 2019: A systematic review and meta-analysis. J Med Virol. 2021;93(2):820–830. doi:10.1002/jmv.26326

22. Buitrago-Garcia D, Egli-Gany D, Counotte MJ, et al. Occurrence and transmission potential of asymptomatic and presymptomatic SARSCoV-2 infections: A living systematic review and meta-analysis. PLoS Med. 2020;17(9). doi:10.1371/journal.pmed.1003346

23. Byambasuren O, Cardona M, Bell K, Clark J, McLaws ML, Glasziou P. Estimating the extent of asymptomatic COVID-19 and its potential for community transmission: Systematic review and meta-analysis. J Assoc Med Microbiol Infect Dis Canada. 2020;5(4):223–234. doi:10.3138/jammi-2020-0030

24. Gombar S, Chang M, Hogan CA, et al. Persistent detection of SARS-CoV-2 RNA in patients and healthcare workers with COVID-19. J Clin Virol. 2020;129. doi:10.1016/j.jcv.2020.104477

25. van Kampen JJA, van de Vijver DAMC, Fraaij PLA, et al. Duration and key determinants of infectious virus shedding in hospitalized patients with coronavirus disease-2019 (COVID-19). Nat Commun. 2021;12(1):267. doi:10.1038/s41467-020-20568-4

26. Walsh KA, Spillane S, Comber L, et al. The duration of infectiousness of individuals infected with SARS-CoV-2. J Infect. 2020;81(6):847–856. doi:10.1016/j.jinf.2020.10.009

27. Stohr Jjjm, Zwart VF, Goderski G, et al. Self-testing for the detection of SARS-CoV-2 infection with rapid antigen tests. medRxiv. Published online February 23, 2021:2021.02.21.21252153. doi:10.1101/2021.02.21.21252153

28. Mcnaughton CD, Adams NM, Johnson CH, Ward MJ, Lasko TA. Diurnal variation in SARS-CoV-2 PCR test results: Test accuracy may vary by time of day. medRxiv. Published online March 13, 2021:2021.03.12.21253015. doi:10.1101/2021.03.12.21253015

29. Minich JJ, Ali F, Marotz C, et al. Feasibility of using alternative swabs and storage solutions for paired SARS-CoV-2 detection and microbiome analysis in the hospital environment. Microbiome. 2021;9(1):25. doi:10.1186/s40168-020-00960-4

30. Medema G, Heijnen L, Elsinga G, Italiaander R, Brouwer A. Presence of SARS-Coronavirus-2 RNA in Sewage and Correlation with Reported COVID-19 Prevalence in the Early Stage of the Epidemic in The Netherlands. Cite This Environ Sci Technol Lett. 2020;7:511–516. doi:10.1021/acs.estlett.0c00357

31. Michael-Kordatou I, Karaolia P, Fatta-Kassinos D. Sewage analysis as a tool for the COVID-19 pandemic response and management: The urgent need for optimised protocols for SARS-CoV-2 detection and quantification. J Environ Chem Eng. 2020;8(5):104306. doi:10.1016/j.jece.2020.104306

32. Nissen K, Krambrich J, Akaberi D, et al. Long-distance airborne dispersal of SARS-CoV-2 in COVID-19 wards. Sci Rep. 2020;10(1):19589. doi:10.1038/s41598-020-76442-2

33. Zhang XS, Duchaine C. SARS-CoV-2 and Health Care Worker Protection in Low-Risk Settings: a Review of Modes of Transmission and a Novel Airborne Model Involving Inhalable Particles. Clin Microbiol Rev. 2020;34(1). doi:10.1128/CMR.00184-20

34. Karimzadeh S, Bhopal R, Huy NT. Review of infective dose, routes of transmission, and outcome of COVID-19 caused by the SARS-COV-2: comparison with other respiratory viruses. Epidemiol Infect. 2021;149:1–22. doi:10.1017/s0950268821000790

35. Sungnak W, Huang N, Bécavin C, et al. SARS-CoV-2 entry factors are highly expressed in nasal epithelial cells together with innate immune genes. Nat Med. 2020;26(5):681–687. doi:10.1038/s41591-020-0868-6

36. Aday S, Aday MS. Impact of COVID-19 on the food supply chain. Food Qual Saf. 2020;4(4):167–180. doi:10.1093/fqsafe/fyaa024

37. Zuber S, Brüssow H. COVID 19: challenges for virologists in the food industry. Microb Biotechnol. 2020;13(6):1689–1701. doi:10.1111/1751-7915.13638

38. Shi J, Wen Z, Zhong G, et al. Susceptibility of ferrets, cats, dogs, and other domesticated animals to SARS-coronavirus 2. Science (80-). 2020;368(6494):1016–1020. doi:10.1126/science.abb7015

39. Mahdy MAA, Younis W, Ewaida Z. An Overview of SARS-CoV-2 and Animal Infection. Front Vet Sci. 2020;7. doi:10.3389/fvets.2020.596391

40. Herstein JJ, Degarege A, Stover D, et al. Characteristics of SARS-CoV-2 Transmission among Meat Processing Workers in Nebraska, USA, and Effectiveness of Risk Mitigation Measures. Emerg Infect Dis. 2021;27(4). doi:10.3201/eid2704.204800

41. Ueki H, Furusawa Y, Iwatsuki-Horimoto K, et al. Effectiveness of Face Masks in Preventing Airborne Transmission of SARS-CoV-2. mSphere. 2020;5(5):2–6. doi:10.1128/msphere.00637-20

42. Cheng Y, Ma N, Witt C, et al. Face masks effectively limit the probability of SARS-CoV-2 transmission. Science. Published online May 20, 2021. doi:10.1126/science.abg6296

43. Lopes H, Middleton J, Martin-Moreno J, et al. Strategic Use of Masks As an Element of a Nonpharmaceutical Measures Set for a Pandemic.; 2020. doi:10.13140/RG.2.2.25214.13125

44. Morawska L, Tang JW, Bahnfleth W, et al. How can airborne transmission of COVID-19 indoors be minimised? Environ Int. 2020;142. doi:10.1016/j.envint.2020.105832

